# Developmental trajectories of child resilience, their determinants, and associations with risk experiences and behaviours: protocol for a national cohort study within the ReDiKid project

**DOI:** 10.64898/2026.10.21.26365702

**Authors:** Jana Pelclová, Sunwoo Lee, Roman Cuberek, Martin Dolejš, Jan Dygrýn, Ivana Hanzlíková, František Chmelík, Jana Kvintová, Roman Procházka, David Smahel, Jaroslava Suchá, Michaela Šaradín Lebedíková, Michał Tkaczyk, Tomas Vetrovsky, Michal Vorlíček, Tomáš Vyhlídal, Aleš Gába

**Author notes:** Correspondence address: Jana Pelclová, Institute of Active Lifestyle, Faculty of Physical Culture, Palacký University Olomouc, třída Míru 117, 771 11 Olomouc, Czech Republic.

## Abstract

**Background:** Resilience during the transition from middle childhood towards early adolescence is best understood as a dynamic, adversity-activated developmental process rather than a fixed individual trait. Building on the *Resilient Kid in the Digital World* framework, this protocol describes the longitudinal component of the broader research programme, designed to characterise developmental trajectories of resilience-related capacities and resources and adaptive functioning, identify their determinants, and examine prospective associations with risk experiences and behaviours across behavioural, family, school, and digital contexts.

**Methods:** This is a national school-based cohort study following children aged 9–11 years at baseline across four waves at approximately six-month intervals from autumn 2025 to spring 2027. Schools are selected from the Czech national register using stratified probability-proportional-to-size sampling by region, municipality status, and area-level socioeconomic status; all eligible Grade 4 and Grade 5 pupils in participating schools are invited. The recruitment plan covers approximately 72 schools and 8,000 eligible children, with an anticipated 50–60% participation rate for complete child–parent pairs, corresponding to a target of approximately 4,000–4,800 linked child–parent dyads at baseline. Longitudinal planning aims to retain at least 1,000 linked dyads through Wave 4. Data are collected through child self-report, parent self- and proxy-report, and school-coordinator online questionnaires. Measures cover resilience-related capacities and resources, adaptive functioning and well-being, risk exposures, digital engagement, health-related behaviours, family context, parental mediation, and school context. Analyses will be tailored to the specific research questions and may include longitudinal and multilevel approaches complemented by reciprocal and person-oriented analyses.

**Discussion:** The study provides a multi-dimensional, nationally distributed longitudinal resource for examining how child functioning and resilience-related resources change together with risk experiences and contextual conditions during a key developmental transition. The design explicitly distinguishes resilience-related measures from evidence of resilience under adversity and treats digital environments as one interacting developmental context rather than as an isolated exposure domain. The cohort may also inform understanding of early vulnerability and protective processes relevant to problematic digital engagement, without treating such indicators as clinical diagnoses.

## Introduction

Middle childhood and the transition towards early adolescence are marked by substantial developmental reorganisation in self-regulation, coping, behavioural autonomy, and social relationships. During this period, direct parental regulation gradually decreases, while peer relationships and school and digital environments become increasingly influential sources of support, norms, and developmental challenge (1–3). These changes make late childhood an important period for examining why children exposed to similar challenges can show different patterns of functioning over time.

The developmental changes occurring during this period are likely to vary considerably across children (4). Resilience-related capacities and resources, together with adaptive functioning, may follow heterogeneous developmental trajectories shaped by individual characteristics and by behavioural, family, school, peer, and digital contexts. Characterising these trajectories and identifying factors associated with their variation is therefore important for understanding heterogeneity in adaptation across the transition to adolescence.

The Resilient Kid in the Digital World (ReDiKid) framework provides the conceptual basis for the present cohort. It defines resilience as a higher-order, adversity-activated process through which adaptive functioning is maintained, restored, or reorganised over time, and situates this process within four interacting domains: psychophysiological, behavioural, contextual, and digital (5). Importantly, resilience is distinguished from the capacities and resources that may support it, from competence, and from adaptive functioning or well-being itself. Consequently, measures of adaptability, social-ecological resources, or well-being are treated in this study as resilience-related capacities, resources, or indicators of functioning; they are not interpreted as direct evidence that a resilience process has occurred unless a relevant adverse exposure and subsequent functioning can be examined in temporal relation.

A longitudinal perspective is required because resources, functioning, and risk exposure may influence one another across development. Individual and behavioural characteristics can shape the situations children subsequently encounter, while family, peer, school, and digital experiences may in turn contribute to later functioning. Digital environments are therefore considered one developmental context among several. They can provide opportunities for learning and social connection as well as exposure to potentially harmful content or interactions, and their associations with well-being vary substantially between children and contexts (6, 7). Family and school resources may also operate as promotive or protective factors across both offline and digitally mediated experiences (8). This transactional perspective provides the rationale for examining both prospective determinants and reciprocal longitudinal associations rather than treating risk exposure or resilience-related capacities as static predictors.

Several gaps motivate the present study. First, longitudinal cohorts beginning in middle childhood that jointly follow resilience-related capacities and resources, adaptive functioning, health-related behaviours, digital experiences, and family and school contexts remain limited. Second, reliance on a single informant can obscure contextual processes and inflate associations through shared method variance; combining child, parent, and school reports provides complementary perspectives (9). Third, average population associations may mask meaningful heterogeneity in how resources and vulnerabilities are configured within children and how these configurations change over time. Finally, resilience research can be conceptually imprecise when resilience scales are treated as direct evidence of successful adaptation without an adversity criterion. The broader ReDiKid programme addresses these challenges through several complemetary study components, each described in dedicated protocol. Within this programme, the present four-wave national cohort has the primary role of mapping medium-term developmental trajectories, their determinants, heterogeneous profiles, and associations with risk experiences and behaviours from middle childhood towards early adolescence. From a developmental psychopathology perspective, these longitudinal patterns may also inform understanding of problematic digital engagement by examining individual vulnerabilities alongside family, peer, school, and digital contexts (10, 11).

### Study Objectives

The present study aims to characterise developmental trajectories of resilience-related capacities and resources and adaptive functioning from middle childhood towards early adolescence, identify individual, behavioural, family, school, and digital determinants of these trajectories, and examine their prospective associations with risk experiences and behaviours.

Specifically, the study addresses five core objectives:

1. To characterise within-child change and between-child heterogeneity in repeatedly measured resilience-related capacities and resources and adaptive-functioning indicators across four waves.
2. To examine prospective associations of individual factors, health-related behaviours, and digital engagement with resilience-related capacities and resources and adaptive functioning.
3. To examine how family and school contexts are prospectively associated with resilience-related capacities, resources, and adaptive functioning and, in prespecified adversity-conditioned analyses, whether contextual resources modify associations between potentially adverse exposures and subsequent functioning.
4. To identify developmental profiles defined by configurations of resilience-related capacities and resources, adaptive functioning, health-related and digital behaviours, risk experiences, and ecological resources.
5. To examine prospective and reciprocal associations between resilience-related indicators and risk experiences or behaviours, including digital risks, and, where exposure meets prespecified proxy adversity criteria, patterns of subsequent adaptive functioning.

## Methods and Analysis

### Study Design

This study is a national, school-based longitudinal cohort study following children aged 9–11 years at baseline in the Czech Republic. A single closed cohort is assessed across four waves over approximately 18 months: baseline (autumn 2025), follow-up 1 (spring 2026), follow-up 2 (autumn 2026), and follow-up 3 (spring 2027), at approximately six-month intervals (Fig. 1). At each wave, the design combines child self-reports completed in school, parent or primary-caregiver self-reports and proxy reports completed remotely, and school-level contextual information reported by designated School Coordinators. This multi-informant structure is intended to capture change within children while also characterising family and school contexts across the transition from middle childhood towards early adolescence.

**Figure 1.**
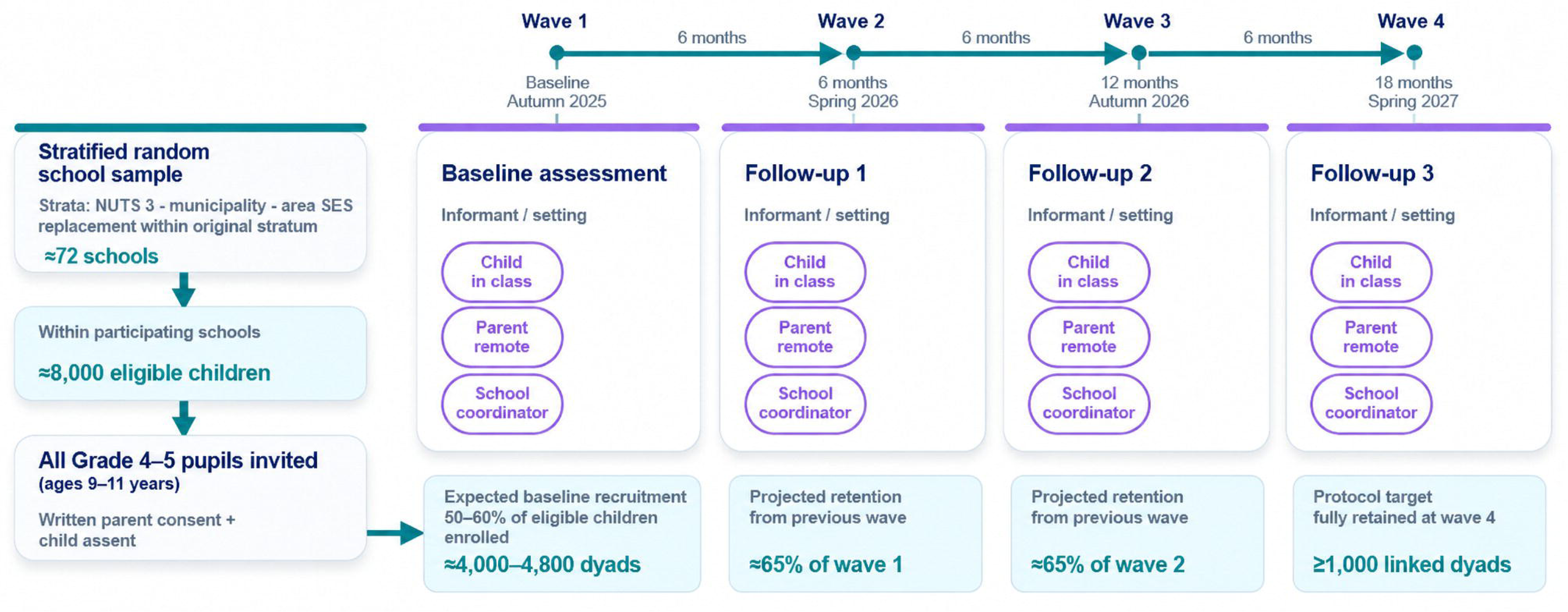
ReDiKid national cohort study design and participant flow. Note. NUTS 3 = Nomenclature of Territorial Units for Statistics, level 3 (regions); SES = socioeconomic status. Stratified school sampling, recruitment planning assumptions, and the four-wave assessment schedule are presented. Numbers of eligible children, targeted baseline parent–child dyads, and projected retention represent planning estimates rather than observed participation. A linked dyad requires questionnaire responses from both the child and a parent or primary caregiver.

### Sampling and Recruitment Procedures

School-based recruitment is used to enrol children and their parents or primary caregivers and to identify designated School Coordinators through a stratified national sample of Czech primary schools.

#### School Selection and Recruitment

The study is conducted in primary schools across the Czech Republic. Schools are selected from the national register of primary schools maintained by the Ministry of Education, Youth and Sports using stratified systematic probability-proportional-to-size sampling. Strata are defined by geographic region (NUTS 3), municipality status (village, town, or statutory city), and area-level socioeconomic status (lower or higher). Area-level socioeconomic status was based on employment rate and the proportion of residents with university education. Both indicators were standardised as *z*-scores and averaged into a composite score, with values <0 classified as lower socioeconomic status and values ≥0 as higher socioeconomic status. The design is intended to achieve broad national coverage across predefined strata; representativeness of the achieved cohort will be evaluated after recruitment and response patterns are known.

The sampling frame includes 4,258 schools, with the school as the sampling unit. Because grade-specific enrolment counts were unavailable in the sampling frame, a measure of size was estimated from school capacity. The target sample is allocated across strata in proportion to the estimated eligible population. Within each stratum, schools are selected systematically with probability-proportional-to-size using a random start and a sampling interval based on the stratum-specific total measure of size and the required number of schools. Larger schools therefore have a higher probability of selection. The allocation is expected to yield approximately 72 sampled schools. If a selected school declines participation, replacement follows the original stratum criteria to preserve the planned sampling structure. The detailed sampling algorithm and stratification procedure are publicly deposited (https://doi.org/10.5281/zenodo.17090817).

Initial contact with school principals uses standardised telephone outreach and formal follow-up communication. Schools receive information on the scientific aims, study procedures, data protection, and expected organisational burden. Participating schools appoint a School Coordinator who serves as the primary liaison across data-collection waves. Coordinators receive protocol training, written guidance, and financial compensation for their organisational role. Recruitment contacts, participation decisions, and reasons for nonparticipation are recorded to document recruitment flow and support evaluation of potential selection bias.

#### Child and Parent Recruitment

Within participating schools, all pupils enrolled in Grades 4 and 5 (approximately 9–11 years) are invited; no within-school subsampling is used. Families receive age-appropriate study information and written consent materials. Participation requires written informed consent from a legal guardian and written child assent at baseline, covering participation across all four waves. No additional exclusion criteria are applied. Stable study identifiers allow linkage of child and parent data and repeated observations while keeping research responses separate from direct identifiers.

#### Target Sample Size and Expected Retention

The recruitment scenario illustrated in Fig. 1 assumes approximately 8,000 eligible children and complete participation of 50–60% of eligible child–parent pairs, corresponding to a baseline target of approximately 4,000–4,800 linked dyads. A dyad is counted as complete only when both the child and the parent or primary caregiver provide questionnaire data. For planning purposes, retention between consecutive waves is projected at approximately 65% (within an anticipated range of 60–70%). The protocol target is at least 1,000 linked dyads at Wave 4.

Potential selection and attrition bias will be evaluated by describing recruitment flow and comparing available baseline demographic, behavioural, and psychosocial characteristics of participants retained at follow-up with those lost to follow-up. Where analyses aim to support population-level inference, design weights and appropriate nonresponse or calibration adjustments will be considered using available sampling and population information.

### Data Collection and Measurement Framework

All study questionnaires are administered through REDCap (Research Electronic Data Capture), hosted on secure servers at Palacký University Olomouc. Stable pseudonymised study identifiers are used to link participants across waves and to link child and parent records while limiting the use of direct identifiers in research datasets.

#### Data Collection Procedures

Data collection spans four waves at approximately six-month intervals. Children complete a self-administered web-based questionnaire during school hours in a classroom or computer-room setting under the organisational supervision of the trained School Coordinator, following standardised instructions provided by the research team. The coordinator may assist with technical orientation or clarification of instructions but is instructed not to influence responses. Parents or primary caregivers receive a secure individualised REDCap link by email and complete the parallel questionnaire remotely.

At each wave, the School Coordinator also receives a secure individualised REDCap link to complete a survey about school policies, practices, and climate. Participants receive unique access codes or links for each wave while longitudinal linkage is maintained through the study identifier system.

#### Follow-up Data Collection and Retention Procedures

Across Waves 2–4, the closed cohort is recontacted in collaboration with participating schools. Before each wave, School Coordinators update participation and school-attendance information and schedule the child survey session. Parents receive secure survey links by email, with systematic reminders used at baseline and follow-up to reduce nonresponse. Participants who withdraw consent or assent are not replaced, and attrition is documented across waves.

#### Measurement Framework

The measurement architecture is informed by the ReDiKid framework (5). For analytical and interpretative purposes, constructs are organised into four functional classes: (1) potentially adverse or risk exposures; (2) individual and contextual capacities and resources that may operate as promotive or protective factors; (3) indicators of adaptive functioning and well-being; and (4) broader individual, behavioural and ecological characteristics that may shape exposure or functioning. These classes are conceptual rather than mutually exclusive. The multidisciplinary battery was designed to balance construct coverage and psychometric quality against respondent burden. Tables 1–3 summarise the child, parent, and School Coordinator questionnaires. Repeated core constructs are retained where longitudinal comparability is required, whereas time-invariant, slowly changing, grade-specific, or long-reference-period modules are administered only in selected waves to limit unnecessary burden. The exact wave- and grade-specific schedule is governed by the locked questionnaire version for each wave; analyses will use only measurements that are comparable for the constructs and time points concerned.

**Table 1.**
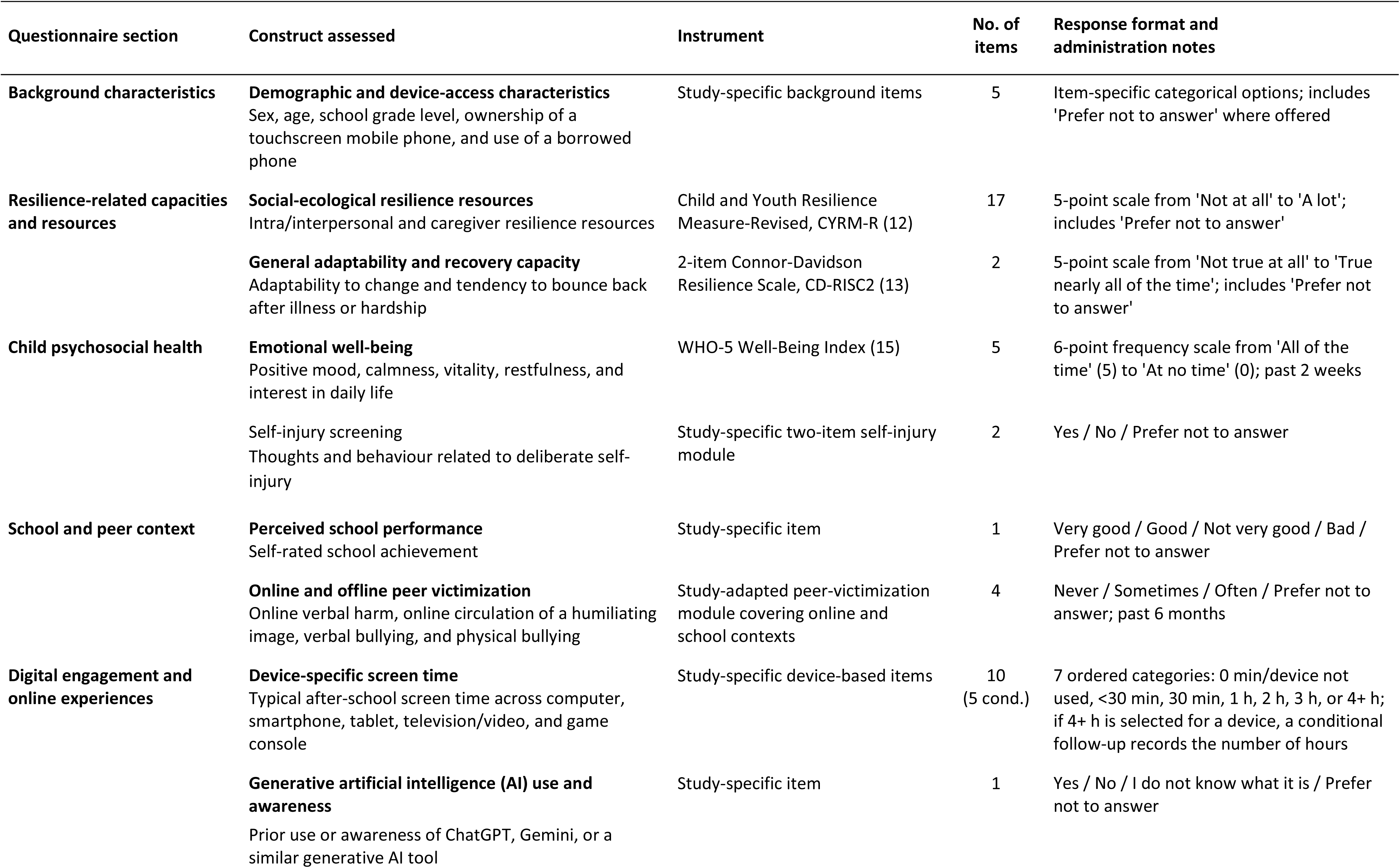
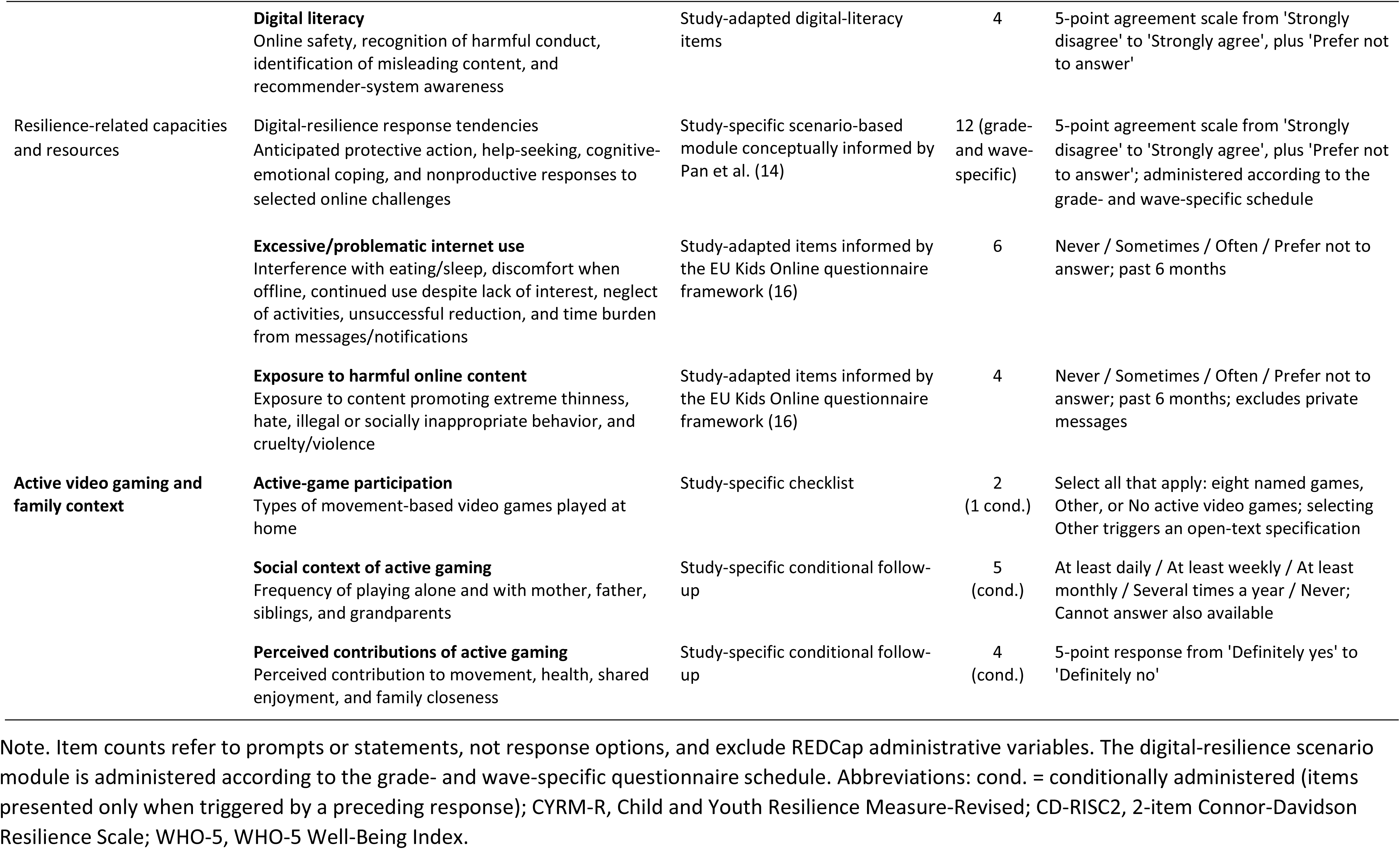
Overview of the Child Self-report Questionnaire (ReDiKid-QCH)

**Table 2.** Overview of the Parent-Report Questionnaire (ReDiKid-QPA)

| Questionnaire section | Construct assessed | Instrument | No. of items | Response format and administration notes |
| --- | --- | --- | --- | --- |
| **Responding parent and household context** | **Parent demographic and anthropometric characteristics**<br>Sex, age, standing height, and body weight | Study-specific background items | 4 | Item-specific categorical and numeric responses; height in cm and weight in kg (weight optional); includes 'Prefer not to answer' where offered |
|  | **Parent socioeconomic and household context**<br>Marital status, education, principal economic activity, cohabiting-partner status, household size and minors, municipality size, and net household income | Study-specific socioeconomic and household items | 8 | Single-choice categorical responses, integer household counts, and an income band; includes 'Prefer not to answer' where offered |
|  | **Cohabiting partner socioeconomic characteristics**<br>Highest completed education and current gainful-employment status of the cohabiting partner/spouse | Study-specific partner-background items | 2 (cond.) | Single-choice categorical responses; asked only when the respondent reports living with a partner/spouse |
| **Child background and health (proxy)** | **Child demographic and anthropometric characteristics**<br>Child sex, date of birth, standing height, and body weight | Study-specific child-background items | 4 | Sex category and date entry (DD-MM-YYYY); height in cm and weight in kg (weight optional); parent proxy-report |
|  | **Child health history, school absence, and educational support**<br>Serious injury or procedure requiring hospitalization/operation and its type and recency; long-term or recurrent health problems and their specification; sickness absence; and recommended educational support measures, including their level and diagnostic or disadvantage basis | Study-specific child-health and education items | 10 (6 cond.) | Four core prompts with six conditional follow-ups; Yes/No and item-specific categorical or open-text responses; integer sickness-absence days (past 6 months); parent proxy-report |
| **Child health-related quality of life (proxy)** | **Health-related quality of life**<br>Physical well-being; psychological well-being; autonomy and parent relations, including leisure-time autonomy, parental time and fairness, parent-child communication, and perceived financial resources; social support and peers; and school environment | KIDSCREEN-27 Parent/Proxy Version (17, 18) | 27 | Five-category global-health item and 5-point intensity or frequency scales, plus 'Prefer not to answer'; past week; parent proxy-report |
| **Parenting and digital mediation** | **Technical parental mediation**<br>Use of software or apps to block websites, limit time or available apps, and restrict the child's online contacts | Study-adapted technical-mediation items informed by EU Kids Online 2020 (16) | 4 | Yes / No for each parental-control function; parent self-report about controls applied to the target child |
|  | **Active and restrictive parental mediation**<br>Encouragement, safety guidance, communication and explanation, time limits, prohibited online activities, and device shut-off rules | Study-adapted parental-mediation items informed by EU Kids Online 2020 (16) | 7 | 5-point applicability/agreement scale from Not at all true to Completely true, plus 'Prefer not to answer'; parent self-report |
| **Child sleep (proxy)** | **Sleep-related difficulties and daytime sleepiness**<br>Unrefreshing sleep, daytime sleepiness, sleepiness observed by a teacher or another supervising adult, and difficulty waking in the morning | Study-specific sleep-related screening items | 4 | Yes / No / Don't know; parent proxy-report |
|  | **Sleep timing**<br>Usual sleep-onset and wake times on school days and weekends | Study-specific sleep-timing items | 4 | Time entry (HH:MM); usual school-week and weekend timing; parent proxy-report |
| **Child physical activity and mobility (proxy)** | **Physical activity participation and volume**<br>Weekly organized and unorganized activity; frequency and type of organized activity and participation in competitions; moderate-to-vigorous physical activity; and muscle-strengthening activity | Study-specific physical-activity items | 7<br>(3 cond.) | Sliders: organized/unorganized activity 0–16 h/week and MVPA 0–10+ h/week; muscle strengthening 0–7 days/week; conditional categorical and open-text follow-ups for organized activity; parent proxy-report |
|  | **Outdoor mobility constraints and perceived safety**<br>Household rules governing the child's unsupervised outdoor time and perceived neighborhood safety for independent mobility | Study-specific outdoor-mobility items | 2 | Item-specific single-choice categorical responses; parent proxy-report |
|  | **Travel to school**<br>Main mode of travel to school and usual journey duration | Study-specific school-travel items | 3<br>(1 cond.) | Single-choice travel mode and 0–120 min slider; selecting Other triggers an open-text specification; parent proxy-report |
| **Active video gaming and family context** | **Active-game participation**<br>Types of movement-based video games played by the responding parent | Study-specific checklist | 2<br>(1 cond.) | Select all that apply: eight named games, Other, or No active video games; selecting Other triggers an open-text specification; parent self-report |
|  | **Active video gaming frequency and family co-play**<br>Frequency of playing active video games alone and with children | Study-specific frequency items | 2<br>(cond.) | At least once daily / weekly / monthly / several times per year / never; separately for playing alone and with children; shown to parents who report playing active video games |
|  | **Perceived contributions of active video gaming to family functioning**<br>Perceived contribution to physical activity, mutual support, healthy habits, shared leisure time, and family bonds | Study-specific perceived-benefit items | 5<br>(cond.) | 5-point response from Definitely yes to Definitely no; parent self-report; shown to parents who report playing active video games |
|  | **Family active video gaming attitudes, intentions, and preferred co-players**<br>Anticipated enjoyment of family co-play, intention to try family co-play, and preferred play context, including playing alone, selected co-players, or not wishing to play | Study-specific conditional follow-up | 4<br>(cond.) | Two 5-point agreement items; multiple-response checklist for preferred co-players; selecting Other triggers an open-text specification; shown to parents who report no active video-game participation |
| **Parent psychosocial resources** | **Parent resilience-related capacity**<br>Perceived adaptability and capacity to cope with and recover from stress or adversity | 10-item Connor-Davidson Resilience Scale, CD-RISC-10 (19) | 10 | 5-point scale from Not true at all to True nearly all of the time; parent self-report |
| **Recent life-event exposure** | **Parent/family life-event exposure**<br>Occurrence of major personal, family, occupational, financial, and lifestyle events during the previous 12 months | Study-adapted life-event checklist based on the Social Readjustment Rating Scale (SRRS) (20) | 43 | Multiple-response checklist: select each event personally experienced during the past 12 months; None of the above also available |
| **Residential environment** | **Housing and neighborhood context**<br>Dwelling type, built-form category, satisfaction with the locality, and municipality and street names | Study-specific residential-context items | 7<br>(2 cond.) | Two single-choice categorical items, each with a conditional open-text specification for Other; locality satisfaction (Yes/No/Prefer not to answer); municipality and street are optional free-text fields |

Note. The ReDiKid-QPA comprises both parent self-report items and parent proxy-report items concerning the child. Item counts refer to prompts, statements, or individual life-event checklist entries rather than response options and exclude REDCap administrative variables. cond. = conditionally administered (items presented only when triggered by a preceding response).

**Table 3.** Overview of the School Coordinator Questionnaire (ReDiKid-QSC)

| Questionnaire section | Construct assessed | Instrument | No. of items | Response format and administration notes |
| --- | --- | --- | --- | --- |
| **Coordinator characteristics** | **Coordinator role and professional background**<br>School role(s), sex, age group, total experience in education or pedagogical work, and tenure at the school | Study-specific coordinator-background items | 6<br>(1 cond.) | Role checklist with a conditional open-text field for Other; single-choice sex, age, and experience categories; includes Prefer not to answer where offered |
| **Study-specific school rules-and-measures matrix** | **Formalization of school rules and measures**<br>Rules or measures concerning healthy eating, physical activity, violence and bullying, netiquette, safe social media use, AI tools, and sexuality education | Study-specific school-policy matrix | 8 | Seven matrix items: not established / established informally / established in writing / don't know; one optional open-text comment; primary level (Grades 1–5) |
|  | **Communication-device rules**<br>Scope and formalization of restrictions on bringing and using communication devices and making audio or video recordings, including other locally applicable rules | Study-specific communication-device rules module | 7 | Five matrix items using the four-category policy scale; two optional open-text fields for other rules and comments; primary level (Grades 1–5) |
|  | **Implementation and enforcement of communication-device rules**<br>Perceived pupil adherence to communication-device rules and frequency of sanctions for rule violations | Study-specific communication-device implementation items | 4 | Two 5-point ratings (adherence and sanction frequency), each followed by an optional open-text comment; past 6 months; primary level (Grades 1–5) |
| **School educational and health-promotion activities** | **Digital safety and digital-skills education**<br>Provision of activities for pupils, professional development for teachers, and parent education, including topic, content, format, and provider | Study-specific digital-education module | 7<br>(3 cond.) | Three Yes/No/Don't know items; each Yes triggers a free-text description; one optional general comment; past 6 months; primary level (Grades 1–5) |
|  | **School support for physical activity**<br>Breaktime and extracurricular activities, movement integrated into lessons, outdoor opportunities, and promotion of active travel to and from school | Study-specific physical-activity support matrix | 6 | Five matrix items using the four-category policy scale, plus one optional open-text comment; primary level (Grades 1–5) |
| **Peer and classroom social climate** | **Peer relationships and mutual support**<br>Overall quality of peer relationships and pupils' willingness to help one another in classes participating in ReDiKid | Study-specific school-climate items | 4 | Two 5-point ratings (very poor to very good; not at all willing to very willing), each followed by an optional open-text comment; past 6 months |
|  | **Pupil fear of classmates and cyberbullying cases addressed**<br>Frequency of situations in which pupils feared classmates and frequency of cyberbullying cases addressed in participating classes | Study-specific peer-risk items | 4 | Never / rarely / sometimes / often / very often; each rating followed by an optional open-text comment; past 6 months |

Note. Item counts refer to individual prompts or matrix rows, including open-text comment fields, rather than response options, and exclude REDCap administrative variables. The School Coordinator questionnaire is administered according to the wave-specific study schedule. cond. = conditionally administered (items presented only when triggered by a preceding response).

#### Child-Reported Measures

The child questionnaire uses standardised response formats, conditional branching, visual section dividers, and age-appropriate administration procedures. Standardised instrument wording is retained. The questionnaire covers the following core domains:

- Background characteristics: Study-specific items capture demographic characteristics, school performance, ownership of a touchscreen mobile phone, and use of a borrowed phone.
- Resilience-related capacities and resources: The 17-item Child and Youth Resilience Measure–Revised (CYRM-R) (12) assesses personal, relational, and caregiver-related social-ecological resources, and the 2-item Connor–Davidson Resilience Scale (CD-RISC2) (13) captures general adaptability and the tendency to recover after hardship. A study-specific scenario-based digital-resilience module, conceptually informed by Pan et al. (14), assesses anticipated protective action, help-seeking, cognitive-emotional coping, and nonproductive responses to selected online challenges in grade- and wave-appropriate administrations. These measures are treated as indicators of capacities, resources, or response tendencies rather than as direct evidence that resilience under experienced adversity has occurred.
- Adaptive functioning and psychosocial health: The WHO-5 Well-Being Index (15) assesses positive mood, calmness, vitality, restfulness, and interest in daily life over the preceding two weeks. A study-specific two-item self-harm screening module asks about thoughts and behaviour related to deliberate self-injury and includes a “Prefer not to answer” option.
- School and peer context: Study-specific and study-adapted items assess perceived school performance and online and offline peer victimisation, including verbal harm, circulation of a humiliating image, verbal bullying, and physical bullying during the preceding six months.
- Digital engagement, literacy, and online experiences: Children report typical device-specific screen time outside school, selected indicators of digital literacy and use of generative artificial-intelligence tools, problematic or excessive internet-use experiences, and exposure to potentially harmful online content. Study-adapted items draw where appropriate on the EU Kids Online questionnaire framework (16).
- Active video gaming and family context: Study-specific items assess children’s participation in movement-based video games at home, types of games played, frequency of playing alone or with family members (mother, father, siblings, and grandparents), and perceived contributions to movement, health, shared enjoyment, and family closeness. Follow-up items are administered conditionally according to reported participation.

#### Parent-Reported Measures

The parent questionnaire combines parent self-reports with parent proxy reports concerning the participating child and covers the following core domains:

- Sociodemographic and residential context: Parent self-report items capture demographic and anthropometric characteristics, education, economic activity, marital and partner status, household composition, income, housing, and residential context.
- Child health and quality of life: Parents complete the KIDSCREEN-27 Parent/Proxy Version (17, 18), covering child’s physical and psychological well-being, autonomy and parent relations, peers and social support, and the school environment domains. Parents also report the child’s demographic and anthropometric characteristics, health history, chronic or recurrent health problems, sickness absence, and recommended educational support.
- Daily routines and health-related behaviours: Parents provide estimates of the child’s screen time and report sleep timing and daytime sleepiness, organised and unorganised physical activity, participation in moderate-to-vigorous and muscle-strengthening activity, travel to school, and aspects of outdoor mobility.
- Active video gaming and family context: Parents report their own participation in movement-based video games and the types of games played. Those who play report the frequency of playing alone or with their children and perceived contributions to physical activity, mutual support, healthy habits, shared leisure time, and family bonds. Parents who do not play report anticipated enjoyment of family co-play, intentions to try it, and preferred co-players.
- Family ecology and parenting: Study-specific items assess family routines and contextual resources, including parental rules, perceived neighbourhood safety, and conditions relevant to the child’s autonomy and everyday functioning.
- Parental digital mediation: Study-adapted items informed by EU Kids Online 2020 (16) assess technical, active, and restrictive mediation, including parental-control functions, safety guidance, communication and explanation, time limits, prohibited online activities, and device shut-off rules.
- Parent psychosocial resources and family stress: Parental resilience-related capacity is assessed using the 10-item Connor–Davidson Resilience Scale (CD-RISC-10) (19). A study-adapted checklist based on the Social Readjustment Rating Scale (20) records major personal, family, occupational, financial, and lifestyle events during the preceding 12 months. These events are treated as indicators of family-level stress or contextual exposure rather than as direct evidence of adversity experienced by the child.

#### School Coordinator-Reported Measures

The School Coordinator Questionnaire is a brief study-specific institutional questionnaire completed by the designated coordinator. It captures the coordinator’s professional background; school rules and measures related to health, behaviour, digital technologies, and communication devices; implementation and enforcement of device rules; digital-safety and digital-skills education for pupils, teachers, and parents; school support for physical activity; and aspects of peer and classroom social climate. Table 3 summarises the constructs and response formats.

### Pilot Testing and Instrument Optimisation

Development and optimisation of the survey battery followed a multistage process to assess developmental suitability and operational feasibility. During the preparatory phase in early 2025, preliminary survey versions were pilot-tested in Czech primary schools that were excluded from the main-study sampling frame.

Pilot work assessed classroom administration time, branching logic, technical functionality, and children’s comprehension. Cognitive interviews with children in the target age range were used to identify ambiguities in study-specific wording and response formats. Findings informed minor structural and linguistic refinements, including visual navigation aids and administration instructions, while the wording of standardised instruments was retained.

### Analytic Strategies

Analyses will be specified in study-specific analysis plans aligned with individual objectives and publications. The common analytical framework recognises repeated measurements nested within children and children clustered within schools, distinguishes within-child change from between-child differences, and avoids treating any single statistical model as mandatory across all outcomes.

#### Descriptive, longitudinal, and multilevel analyses

Baseline and wave-specific characteristics, item nonresponse, recruitment flow, and attrition will be described at child, parent, and school levels. Depending on the outcome and research question, longitudinal change may be examined using mixed-effects, growth-curve, or other repeated-measures models that accommodate the distribution and measurement properties of the construct. School-level clustering and relevant individual- and contextual-level determinants will be incorporated where appropriate. For repeated latent constructs, measurement comparability across time will be evaluated before substantive change is interpreted. Sampling weights and, where appropriate, nonresponse or calibration adjustments will be incorporated in analyses intended to support population-level inference.

#### Reciprocal, contextual, and person-oriented analyses

Where constructs are measured comparably over sufficient waves, prospective and reciprocal models may be used to examine temporal associations between prior functioning or resilience-related indicators and subsequent risk experiences or behaviours. Contextual moderation analyses will evaluate whether family or school resources alter associations between risk exposure and later functioning when measurement timing supports such inference. Person-oriented approaches may be used to identify subgroups characterised by distinct configurations or trajectories of resources, functioning, behaviours, and risk experiences when the data structure and sample size support reliable estimation.

#### Adversity-conditioned analyses

Analyses will be restricted to observations or subgroups meeting prespecified proxy criteria for potentially adverse exposure using available digital, peer, social, family, or other relevant indicators. Meeting a proxy criterion will be treated as an analytical operationalisation of potentially adverse exposure, not as proof that every exposed child experienced the event as psychologically adverse. Operational definitions, thresholds, timing, and corresponding adaptive-functioning outcomes will be prespecified before outcome analyses and documented in the relevant analysis plan. Where temporal ordering is supported, these analyses will examine whether antecedent individual or contextual resources are associated with maintained, restored, or comparatively favourable subsequent functioning.

#### Missing data and statistical reporting

Longitudinal analyses will use all available observations where the selected method permits, rather than relying exclusively on fully retained dyads. Missingness and attrition patterns will be examined, and model-appropriate approaches to missing data will be used under clearly stated assumptions, with sensitivity analyses where feasible. Effect estimates and uncertainty intervals will be prioritised alongside p-values, and causal language will be avoided for observational associations unless justified by the design and analysis.

### Ethical Considerations

The overarching ReDiKid project received ethical approval from the Ethics Committee of the Faculty of Physical Culture, Palacký University Olomouc (Approval No. 41/2024, 19 March 2024). The present longitudinal study subsequently received specific approval from the same committee (Approval No. 10/2026, 13 January 2026). The study is conducted in accordance with the Declaration of Helsinki, applicable Czech legislation, and Regulation (EU) 2016/679 (General Data Protection Regulation; GDPR).

Participation is voluntary and requires written informed consent from a legal guardian together with child assent. Children are informed that the questionnaire is not a test, may skip any question, and may stop participation without negative consequences; guardians may withdraw consent at any time. Families completing all four waves receive CZK 1,500 as compensation for the time associated with repeated participation; compensation is not linked to the content of responses. School Coordinators are trained to support privacy and neutral administration. If a child spontaneously communicates distress or a potential safety concern during data collection, the coordinator follows the school’s established support procedures; questionnaire responses are not used by the research team for individual clinical monitoring.

### Data Security and Pseudonymisation

Research questionnaires are collected and stored in REDCap hosted on secure university infrastructure at Palacký University Olomouc. Direct identifying and contact information required for consent and longitudinal administration is kept separate from research-response data, with linkage performed through pseudonymised study identifiers. Access is restricted to authorised project personnel according to role.

Analytical exports are stripped of direct identifiers. Any data prepared for sharing will undergo additional disclosure-risk assessment and de-identification; variables that cannot be released safely will remain under controlled access in accordance with project governance and the Data Access Committee.

## Discussion

### Conceptual contribution

A central contribution of this cohort is the operationalisation of the ReDiKid framework in a medium-term longitudinal design. Digital environments are treated as a distinct but interacting developmental domain rather than as an isolated or inherently harmful exposure. The study can therefore examine how digitally mediated risks and opportunities co-occur with behavioural, family, school, and individual resources, and whether contextual resources are associated with more favourable functioning following potentially adverse exposures (5).

The protocol also preserves the framework’s distinction between resilience-related capacities and resources, adaptive functioning, and resilience as an adversity-activated process. Repeated questionnaire scores are not used to label children globally as “resilient” or “non-resilient”. Instead, cohort-wide analyses describe developmental trajectories of capacities, resources, functioning, and exposure, while resilience-process interpretations are reserved for adversity-conditioned analyses. Adaptive functioning is likewise treated as domain-specific: favourable functioning in one developmental domain may coexist with difficulties in another. These findings may also inform developmental psychopathology by identifying early vulnerabilities and protective resources associated with problematic digital engagement, without equating such indicators with behavioural addiction (10, 11).

### Methodological strengths

The four-wave design allows within-child change and temporal ordering to be examined across the transition from middle childhood towards early adolescence, while the stratified probability-proportional-to-size school sample provides broad national coverage. The multi-informant structure combines children’s own reports with parent and school information, reducing reliance on a single perspective and enabling contextual interpretation of child-level findings (9). The measurement battery combines established instruments with study-specific measures needed to capture emerging digital and school-policy constructs.

A further strength is analytical flexibility. The cohort can support variable-centred longitudinal analyses as well as person-oriented approaches that identify subgroups with distinct combinations of resources, functioning, behaviours, and exposures. This is important because average associations may conceal developmental heterogeneity and potentially relevant intervention targets.

### Limitations and practical considerations

The ReDiKid longitudinal study is observational; temporal ordering and within-person analyses can strengthen developmental inference but do not by themselves establish causality. Participation also depends on school cooperation, guardian consent, and continued engagement across waves. The achieved cohort may therefore differ from the target population despite the probability-based sampling design. Recruitment, response, and attrition patterns will need to be considered when interpreting generalisability.

Many constructs rely on child or parent self-report and proxy-report and are consequently susceptible to recall error, social desirability, and informant-specific perspectives. The multi-informant design reduces dependence on a single reporter but does not eliminate measurement error; disagreement between informants may itself reflect meaningful differences in perspective or opportunity for observation.

Chronological age and school grade do not fully capture individual developmental timing. Children of similar age may differ in maturational and psychosocial development, so some heterogeneity attributed to wave or age may also reflect unmeasured differences in developmental stage. In addition, although six-month spacing is appropriate for medium-term developmental change, this design cannot capture acute regulation or short-term recovery following specific events.

Finally, available risk indicators do not establish perceived threat, functional disruption, or recovery for every reported exposure. Risk exposure will therefore not automatically be labelled as adversity. Interpretations in terms of resilience processes will be restricted to analyses with prespecified proxy adversity criteria and temporally subsequent adaptive-functioning outcomes, and those outcomes will be interpreted within their specific developmental domains.

## Conclusion

By following a national school-based cohort, this study will characterise developmental trajectories of resilience-related capacities and resources and adaptive functioning, identify their individual, behavioural, and contextual determinants, and examine how they interrelate with risk experiences and behaviours from middle childhood towards early adolescence. The study’s conceptual distinction between resilience-related indicators and resilience under adversity, together with its multi-informant and multilevel design, is intended to support more precise and context-sensitive evidence on children’s development across interconnected offline and digital environments.

## Availability of data and materials

Data collection is ongoing. Following completion of the study, de-identified analytical datasets, data dictionaries, and analytical code will be made available where appropriate and legally permissible, subject to disclosure-risk assessment, project governance, and Data Access Committee procedures. Sensitive or potentially identifying variables may be available only under controlled access.

## Authors’ contributions

JP: Conceptualization, Methodology, Project administration, Supervision, Writing – original draft, Writing – review & editing.

SL: Writing – original draft, Writing – review & editing.

RC: Conceptualization, Methodology, Visualization, Writing – original draft, Writing – review & editing. AG: Conceptualization, Methodology, Writing – review & editing.

MD, JD, IH, FC, JK, RP, DS, JS, MŠL, MT, TV, MV, and TVy: Methodology, Writing – review & editing. All authors critically reviewed the manuscript and approved the final version.

## Acknowledgements

The authors are grateful to the children and their parents or primary caregivers for their participation and commitment to the longitudinal study. We sincerely thank the participating schools, their leadership, teachers, and School Coordinators for facilitating recruitment and supporting data collection across study waves. We also acknowledge the wider ReDiKid research and administrative team for their contributions to pilot testing, fieldwork, technical implementation, and data management.

